# Allergic asthma and type-2 immunity reduce COVID-19 severity

**DOI:** 10.1101/2025.06.16.25329662

**Authors:** Ming Zheng

**Author notes:** Web of Science-ResearcherID: http://www.webofscience.com/wos/author/record/AAM-8056-2021. National Natural Science Foundation of China -Basic Researcher ID: https://cstr.cn/BRID-07816.00.51810. Corresponding authors: **Ming Zheng**, MD, PhD, Beijing Institute of Basic Medical Sciences, 27 Taiping Road, Beijing 100850, China (E-mail address or).

## Abstract

The COVID-19 pandemic, caused by severe acute respiratory syndrome coronavirus 2 (SARS-CoV-2), has had a profound global impact, resulting in millions of cases and deaths. As COVID-19 continues to circulate, the interplay between COVID-19 and other chronic diseases, particularly asthma, remains an important public health concern. Asthma, a heterogeneous disease affecting over 300 million people globally, is characterized by airway inflammation and hyper-reactivity. Asthma exacerbations are often triggered by respiratory infections, leading to an increased risk of pulmonary complications. Given that the COVID-19 pandemic has spurred significant debate on its relationship with asthma, understanding how different asthma phenotypes, particularly allergic versus non-allergic asthma, affect COVID-19 risk is critical. Using Mendelian randomization (MR), this study investigates the genetic determinants of asthma subtypes in relation to COVID-19 susceptibility and severity. Our findings reveal a complex relationship, with non-allergic asthma associated with increased COVID-19 risk, while allergic asthma, particularly when co-occurring with allergic rhinitis, correlates with reduced COVID-19 risk. Although COVID-19 and allergic asthma are independent clinical manifestations, the above results indicate the trade-off between allergic asthma and COVID-19 risk, implying a shared origin of allergic disease and allergic immune defense. As such, I proposed an alternative perspective for allergic diseases: the ability to mount an allergic immune defense can be protective in the face of infectious diseases such as COVID-19; however, when excessive and misguided to allergens, the allergic immune defense can result in allergic disease. Thus, the dramatic rise of allergic diseases during the past few decades might be caused by the survival advantage of allergic immune defenses, which increases the adaptation of the host to lethal infections and thus is favored in the evolutionary process of natural selection.

**Highlights:** COVID-19 pandemic complicates disease progression and management in the asthmatic population.

COVID-19 risk increases with non-allergic asthma but decreases with allergic asthma.

The trade-off between allergic asthma and COVID-19 risk implies a shared origin of allergic disease and allergic immune defense.

## Introduction

Asthma, characterized by airway inflammation and airway hyper-reactivity, is a major public health problem.^1^ Asthma affects approximately 300 million people worldwide, and its prevalence has increased dramatically since the 1950s in developed countries.^1,2^ As a very common but heterogeneous disease, the pathogenesis of asthma involves complex environmental factors, different molecular pathways, many cell types and cytokines.^3^ Although animal models have helped to clarify distinct forms of asthma phenotypes—the classical paradigm of allergic and non-allergic asthma,^4–7^ there is still a lack of human data to integrate heterogenous asthma phenotypes into the broader canvas of complex environmental exposures.^8^ For example, asthma symptoms can be precipitated by viral respiratory infections, regardless of the presence of allergens.^9,10^ The co-occurrence of respiratory infections and asthma contribute to nearly 80% of pulmonary disease exacerbations in both children and adults.^11,12^ This finding leads to an important question: is asthma a friend or foe for the respiratory viral infections? This puzzle has become increasingly important with the advent of post-COVID-19 era.^13,14^

After five years into the coronavirus disease 2019 (COVID-19) pandemic, COVID-19 has turned endemic like the common cold.^15^ Since the beginning of the COVID-19 pandemic, the relationship between asthma and COVID-19 risk has become a hotly debated issue in the field. Different studies of COVID-19 have reported varying prevalence of asthma.^16^ In a meta-analysis of 150 studies worldwide that examined the prevalence and severity of COVID-19 in patients with asthma, there was no consistent relationship between asthma and COVID-19 diagnosis or severity, indicating the complexity of this issue.^17^ Asthma is a heterogeneous disease comprising distinct clinical subtypes with different underlying etiologies. As such, aetiological factors unique to different asthma subtypes (allergic and non-allergic asthma) cannot be excluded. However, such heterogeneity was not investigated in prior research. Thus, the previous results cannot be unequivocally viewed as reflecting the phenotypes of distinct asthma subtypes but must be interpreted in the context of the complexity of asthmatic disease. Such heterogeneity, coupled with multiple confounding factors, creates a complex puzzle that has stymied investigators and clinicians, so there is still a limited understanding of the direct inference between asthma and COVID-19. Understanding this issue will improve preparedness for the recurring waves of COVID-19 ahead and help reduce the overall burden of COVID-19 worldwide.

## Material and Methods

### Study design and data sources

The heterogeneity of asthma and the diversity of potential confounders may overwhelm the ability to study COVID-19 risk in patients with asthma. This problem in conventional observational studies can be overcome by modeling causal inference through the use of Mendelian randomization (MR) method **(As shown in Fig. 1)**. The MR method uses genetic variants, which remain unchanged throughout an individual’s lifetime, to bypass potential confounding factors and are thus less susceptible to confounding bias or reverse causation.^18,19^ Thus, this study performed a two-sample MR analysis to infer the causal effect of genetically-predicted asthma sub-phenotypes on COVID-19 susceptibility and severity.

**Figure 1.**
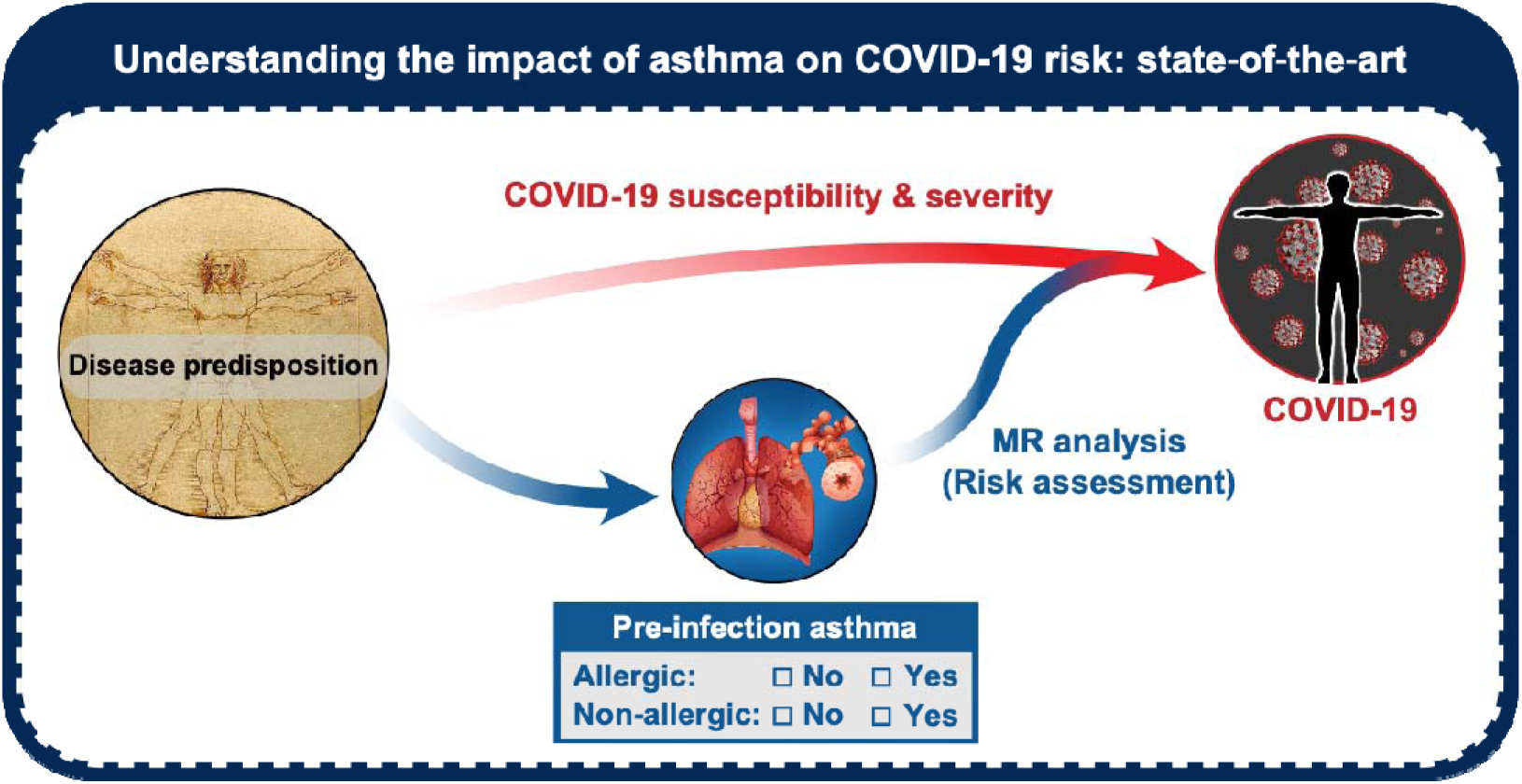
Understanding the impact of asthma on COVID-19: state□of□the□art. There is a huge gap in understanding the impact of asthma on COVID-19. Using Mendelian randomization (MR) analysis for risk assessment, this study investigated the causal inference between pre-infection asthma and the risk of COVID-19 susceptibility and severity. Additionally, this study demonstrates a research workflow that performs the translation of the pre-infection information of asthma into a potential indicator of future COVID-19 risk.

All FinnGen phenotypes were defined from nationwide electronic health-record codes (ICD-8-10, procedure codes, medication reimbursement) and curated by the FinnGen Clinical Endpoints Committee. Pan-UKBB phenotypes followed the PheCode mappings supplied with the Pan-UKBB v1.0 manifest. For allergic versus non-allergic asthma, individuals with any physician-diagnosed allergic disease (allergic rhinitis, atopic dermatitis, food allergy, urticaria) were classified as “allergic”; those without were “non-allergic”.

### Selection of genetic instruments and the exposure and outcome data

Given the basic principle of MR method described above, this study used the genome-wide association study (GWAS) data of asthma from FinnGen and UK biobank, and genetic instruments of single nucleotide polymorphisms (SNPs) for different asthma phenotypes were chosen using a *p* value <5×10^-7^ and an independent inheritance with a minimal level of linkage disequilibrium (LD) of r^2^ <0.001. Instrumental strength was measured using *F*-statistic >10 as sufficiently informative. A total of 290 SNPs were selected with an average *F*-statistic of 53.2 (range 25.3 to 470.2).

### Mendelian randomization analysis

MR analysis was performed to estimate the impact of asthma (exposure) on COVID-19 susceptibility and severity (outcome). To account for potential pleiotropy, we applied the MR pleiotropy residual sum and outlier (MR-PRESSO) test, adjusting for horizontal pleiotropy where necessary using default settings. This approach allowed us to disentangle the causal effects of different asthma phenotypes on COVID-19 risk while minimizing confounding influences.

### Software for statistical analysis

All analyses were conducted in R 3.6.1 using the packages TwoSampleMR (v0.5.6) for core MR methods, MRPRESSO (v1.0) for pleiotropy correction, and ld_clump in ieugwasr (v0.1.5) for LD clumping on SNP instruments. LD reference used the 1000 Genomes Phase 3 European panel.

## Results

### COVID-19 risk increases with non-allergic asthma but decreases with allergic asthma

MR analysis was performed to dissect the specific effect of single asthma subtypes on COVID-19 risk. By interrogating the GWAS data from COVID-19 Host Genetics Initiative,^20^ this MR study analyzed genetically determined asthma in a COVID-19 cohort of 1,683,768 participants. The risk of COVID-19 susceptibility and severity (outcome) was estimated following different asthma subtypes (exposure) **(Fig. 2A)**.

**Figure 2.**
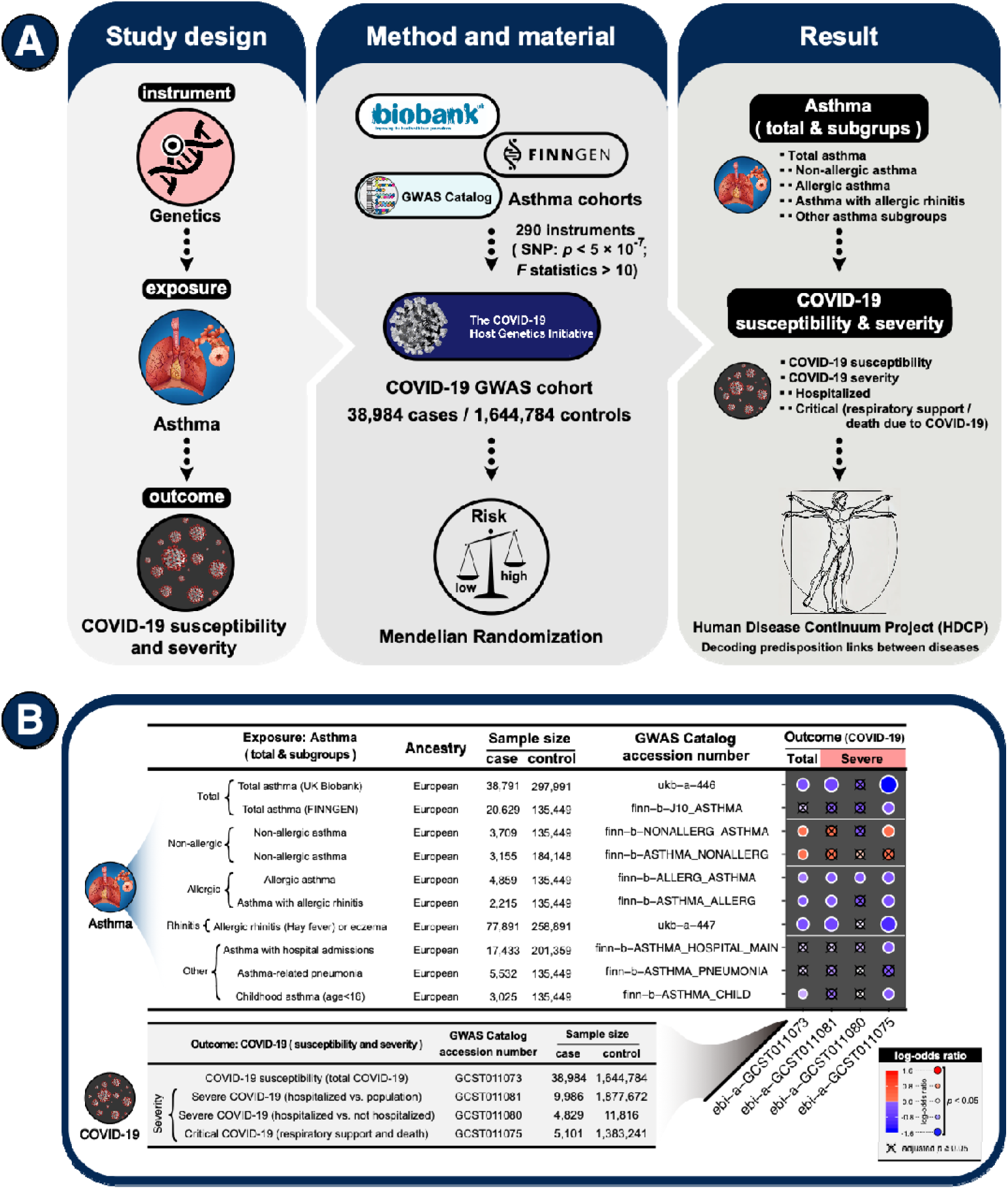
Mendelian randomization study of asthma and COVID-19. **(A)** The graphical illustration of the Mendelian randomization (MR) study within the framework of the Human Disease Continuum Project (HDCP)—an initiative that seeks to decode predisposition links between pathogen exposure and chronic disease through large-scale immune-profiling and longitudinal genomics. The MR analysis was conducted using genetic instruments to estimate the causal role of asthma exposure in COVID-19 susceptibility and severity. This MR analysis investigated the 290 genetic variants of asthma against COVID-19 susceptibility and severity in genome-wide association study (GWAS) cohorts to evaluate the risk of COVID-19 following asthma. **(B)** MR results show the causal effect of asthma exposure on the outcomes of COVID-19. MR estimated causal effects were calculated using the MR pleiotropy residual sum and outlier (MR-PRESSO) test, with horizontal pleiotropy corrected using default settings. The causal estimates were shown by heatmap, with the dot color and size representing the log-odds ratio. A positive log-odds ratio indicates that asthma exposure is associated with an increased risk of COVID-19, and vice versa. The “×” symbol represents false discovery rate (FDR) adjusted p-value ≥ 0.05.

As shown in **Fig. 2B**, asthma exposure was significantly associated with the risks of COVID-19 susceptibility and severity. Significantly decreased COVID-19 risks were found in total asthma (log-odds ratio = -0.21 to -1.63; FDR-adjusted *P*_MR-PRESSO_ <0.05) and the subtypes of allergic asthma (log-odds ratio = -0.04 to -1.04; FDR-adjusted *P*_MR-PRESSO_ <0.05), asthma with allergic rhinitis (log-odds ratio = -0.06 to -1.04; FDR-adjusted *P*_MR-PRESSO_ <0.05), asthma with hospital admissions (log-odds ratio = -0.15; FDR-adjusted *P*_MR-PRESSO_ <0.05), and childhood asthma (log-odds ratio = -0.03 to -0.14; FDR-adjusted *P*_MR-PRESSO_ <0.05). Comparatively, significantly increased COVID-19 risks were found in non-allergic asthma (log-odds ratio = 0.07 to 0.22; FDR-adjusted *P*_MR-PRESSO_ <0.05).

Using MR analysis for risk assessment, this study demonstrates a research workflow that performs the translation of the pre-infection information of asthma into a potential indicator of future COVID-19 risk. For the purpose of discussion, this perspective focuses on two major classes of asthma—allergic and non-allergic asthma. In summary, COVID-19 risk increases with non-allergic asthma but decreases with allergic asthma. The allergic and non-allergic forms of asthma may serve as a binary indicator of COVID-19 risk (**Fig. 1**).

Apart from the classical paradigm of allergic and non-allergic asthma, this study also revealed that other asthma subtypes, including severe asthma (with hospital admissions) and childhood asthma (age < 16 years), are also associated with decreased COVID-19 risk. Compared to allergic and non-allergic asthma using different pathogenic mechanisms that are dependent on or independent of T helper 2 (Th2) cells,^4–7^ an alternative pathogenic mechanism of activated alveolar macrophages interacting with natural killer T cells was found in patients with severe asthma ^9^ and in children with asthma undergoing viral infection.^10^ It has become increasingly apparent that heterogeneous asthma subtypes and the distinct clinical forms or phenotypes reflect different pathogenic mechanisms and biological pathways. Additionally, although different asthma subtypes can develop independently of each other, the underlying pathophysiological pathway may interact and share with each other. To better understand their impact on COVID-19 risk, the classic paradigm of asthma should be extended to include additional nontraditional subtypes.

This open field of research requires more studies. Future research on asthma should be synthesized with the goal of disentangling and understanding the impact of different asthmatic pathways on host defense against respiratory infections. Such studies will probably lead to novel therapies for COVID-19 that should be incorporated into precision medicine, with the different pathogenic mechanisms that occur in each asthma subtype taken into account for individualized and personalized therapy.

### Allergic immune defense against COVID-19 infection

It is well-acknowledged that the co-occurrence of respiratory infections and asthma contributes to disease exacerbations. In our analysis of COVID-19 infection, this common sense was met with the finding that non-allergic asthma increased COVID-19 risk. Strikingly, apart from non-allergic asthma, many of the asthma subtypes—mostly allergic asthma—have a significant association with decreased risks of COVID-19 infections (**Fig. 2B; Supplementary Fig. 1**). Of note, the most robust causality of decreased COVID-19 risk was found in asthma with allergic rhinitis. Allergic rhinitis was significantly associated with different risks of COVID-19, ranging from COVID-19 susceptibility, severe COVID-19, and critical COVID-19 that required respiratory support or caused death. This finding was validated in another cohort of allergic rhinitis (ukb-a-447; **Fig. 2B**). Asthma and allergic rhinitis often coexist: up to 40% of allergic rhinitis patients have asthmatic symptoms, and up to 80% of asthma patients have symptomatic allergic rhinitis.^21^ Thus, the above finding indicates that the allergic phenotype (rhinitis) of asthma may play a vital role.

The above observation suggests that the protective effect of allergic asthma (rhinitis) could exert itself by reducing COVID-19 susceptibility and preventing the progression of the disease. Allergic asthma is mediated by an allergic response of immune system to allergens, which relies on several defensive mechanisms that should provide adequate protection against infections. First, the barrier defense is provided by prolonged or excessive mucus production—a common symptom of allergic asthma (rhinitis). This basic defensive strategy can prevent or minimize the settlement of viruses at the mucosal surfaces and entry into internal compartments. Second, the removal or expulsion defense is characterized by several symptoms, such as sneezing and coughing, that ensure the avoidance of dangerous environmental exposures.

The above-mentioned mechanisms appear to help reduce the risk of COVID-19 infection. However, it should be noted that COVID-19 predisposition may not be fully accounted for by such basic defensive strategies alone. The protective effects of allergic response against COVID-19 infection are likely to depend on a complex mixture of barrier defense, expulsion, and immune mechanisms.

To investigate the underlying mechanisms, the analysis of the genetic variants showed a negative correlation between the SNP effects on allergic asthma and COVID-19 risk (p = 0.038, Spearman’s rho = -0.457; **Fig. 3A**). The most relevant SNPs of allergic asthma, rs74630264 (*IL4R*: Interleukin 4 Receptor) and rs116236597 (*TNFSF4*: Tumor Necrosis Factor Ligand Superfamily Member 4), showed reverse impacts on COIVD-19 risk (**Fig. 3B-C**). The TNFSF4 functions in T cell antigen-presenting cell interactions and mediates adhesion of activated T cells to endothelial cells; the IL4R can bind interleukin 4 and interleukin 13 to regulate IgE production and promote differentiation of Th2 cells. Thus, it appears that the protective effect against COVID-19 infection requires type-2 immune responses, implying the important role of allergic immune defense against respiratory infection.

**Figure 3.**
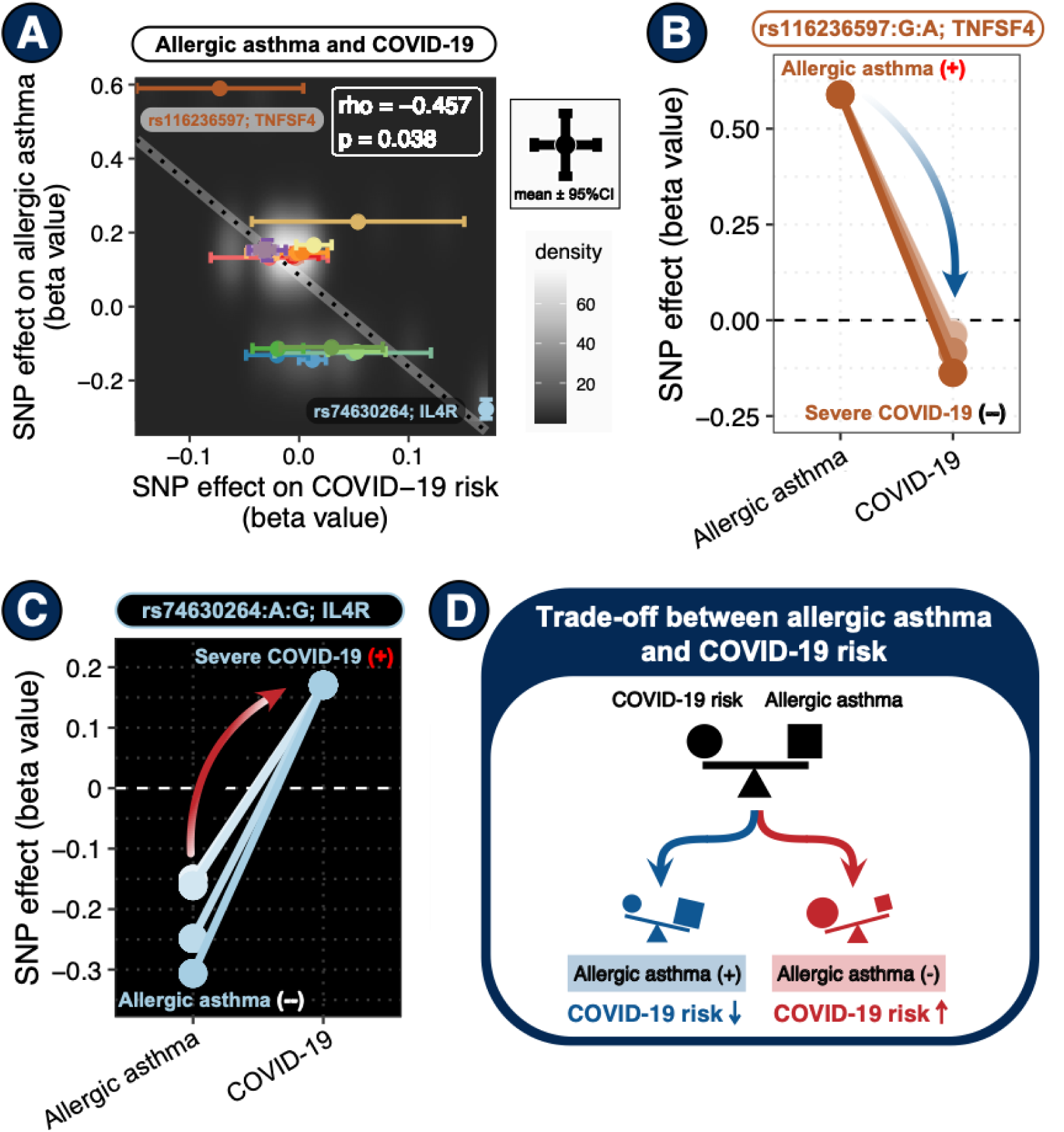
Effects of genetic variants on allergic asthma and COVID-19 risk. **(A)** The correlations between the SNP effects (beta value) on allergic asthma and COVID-19 risk. Dot colors correspond to different genetic variants of SNPs, with error bars representing 95% confidence interval (CI). Spearman’s rho coefficient with p value and linear regression line are indicated in the graph. **(B-C)** The SNP effects (beta value) of rs74630264 (*IL4R*: Interleukin 4 Receptor) and rs116236597 (*TNFSF4*: Tumor Necrosis Factor Ligand Superfamily Member 4) on COVID-19 risk in different clinical subtypes of asthma. **(D)** The graphical illustration shows the trade-off between allergic asthma and COVID-19 risk.

It is worth noting that COVID-19 is a novel disease caused by the emerging SARS-CoV-2 virus since 2019. Although COVID-19 and allergic asthma are independent clinical manifestations, the above results indicate the trade-off between allergic asthma and COVID-19 risk (**Fig. 3D**). This trade-off, reflected by the double-edged effects of allergic immune defense, embodies a fundamental principle of Chinese philosophy—the theory of *Yin* (阴; unfavorable effect) and *Yang* (阳; favorable effect). The *Yang* (bright) side of allergic immunity reduces the risk of COVID-19, whereas the *Yin* (dark) side of allergic immunity increases the risk of allergic asthma. As in the Yin-Yang philosophy, since the *Yang* side of allergic immunity is important in fighting viral infection, considering that it is interconnected with its *Yin* side—allergic disease, this opposite but interconnected Yin-Yang relationship can be omnipresent in any individual and determines disease susceptibility to both allergy and COVID-19, thus explains the existence of allergic diseases in modern humans.

## Discussion

Primary prevention and early treatment of COVID-19 are of utmost importance for reducing COVID-19 mortality. Therefore, finding those with higher COVID-19 risk has become increasingly critical. To our knowledge, this study first sought to confirm the causal impact of asthma on COVID-19 susceptibility and severity, providing convincing evidence at the genetic level. Given the causal inference between COVID-19 and asthma, asthma could be a natural pre-infection condition to predict the future risk of COVID-19. Of course, the above findings require further validation, and future COVID-19 research should also take the difference between allergic and non-allergic asthma into account.

According to this study, an allergic response can provide effective protection against viral infection. Given this finding, the allergic response can be beneficial and not simply a noxious overreaction to allergens. Here, I proposed a relaxing hypothesis that the inborn allergic predisposition could contribute to both allergic disease and immune defense. From the perspective of natural selection, allergic disease could be an acceptable cost of the immune defense against severe and life-threatening infections, herein termed “allergic immune defense.” Allergic immune defense increases the adaptation of the host to lethal infections and thus is under positive selection that enables the passing of this adaptation to offspring. It is reasonable to assume that this adaptation designed for an ancient environment of high pathogen burden could become maladaptive in the super-hygiene environment of modern society.

In conclusion, there is a shared origin of allergic disease and allergic immune defense, despite these being independent clinical manifestations. Based on the above hypothesis, the high prevalence of allergic disease in humans may reflect an important role of allergic immune defense against severe and life-threatening epidemics and pandemics, while allergic disease could be a noxious result of overreaction when allergic immune defense is mistargeted to allergens. An analogous example is that the adaptive immune system evolved to protect from potential environmental threats, but it can result in autoimmune disease when adaptive immunity is mistargeted to self-antigens.^14^

The above-mentioned examples indicate the trade-offs between the benefit of host survival (immune responses to environmental threats) and the cost of noxious overreactions (allergic and autoimmune diseases). Since host survival is always the top priority, there should be a biological or evolutionary advantage for the allergic immune defense to exist in humans. Thus, the genetic predisposition of allergic immune response can be favored in the process of natural selection. A previous study observed a positive selection on the genetic variants of Th2 immune response pathway,^22^ implying that recent evolutionary events may exert a selective pressure that leads to the elevated immune responses at the expense of the increased susceptibility to allergic asthma. This trade-off, reflected by the double-edged effects of allergic immune defense, which may help explain the existence of allergic diseases in modern humans. Thus, the dramatic rise of allergic diseases during the past few decades might be caused by the survival advantage of allergic immune defenses which is favored by the evolutionary process of natural selection (**Fig. 4**).

**Figure 4.**
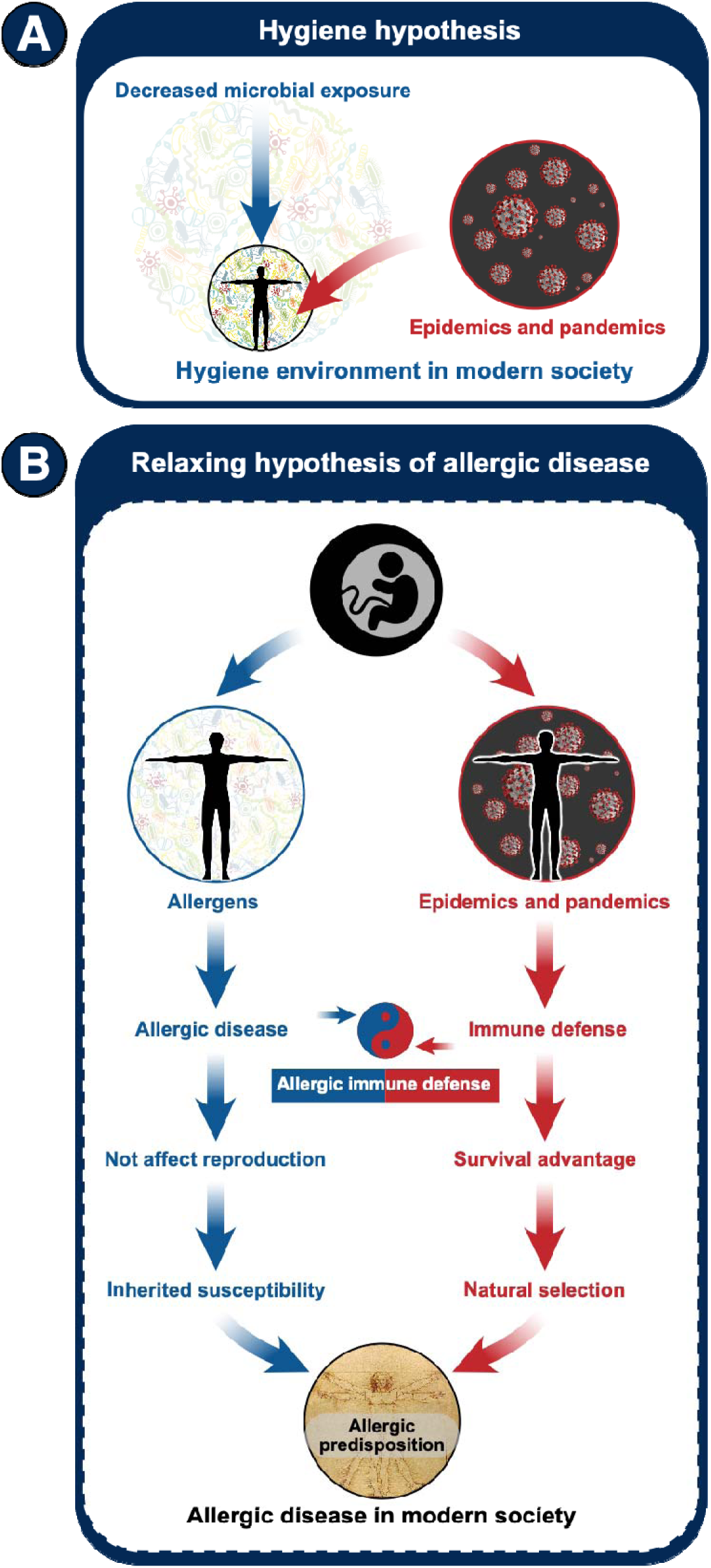
Hygiene hypothesis and allergic disease. **(A)** Hygiene hypothesis. The environments of modern society are characterized by decreased environmental pathogen burden. However, large and highly-connected modern society is particularly vulnerable to epidemics and pandemics. Understanding this hypothesis can help explain why modern environments may cause human diseases. **(B)** Relaxing hypothesis of allergic disease in the context of modern environments. The inborn allergic predisposition could contribute to both allergic disease and immune defense. From the perspective of natural selection, allergic disease could be an acceptable cost of the immune defense against severe and life-threatening infections, herein termed “allergic immune defense.” When excessive and misguided to allergens, the allergic immune defense can result in allergic disease. The double-edged effects of allergic immune defense embody a fundamental principle of Chinese philosophy—the theory of yin (unfavorable effect) and yang (favorable effect), indicating the trade-off between the benefit of host survival and the cost of noxious overreactions, where host survival is the top priority that is always favored by natural selection.

Within the emerging framework of the Human Disease Continuum Project (HDCP)—an initiative that seeks to decode the molecular and immunological mechanisms linking pathogen exposure to chronic disease susceptibility through large-scale immune-profiling and longitudinal genomics^13,14,23–25^—this study on asthma sub-phenotypes extend the disease-continuum concept beyond autoimmunity into the field of allergic disorders. By showing that the very alleles that license a vigorous allergic immune defense simultaneously lower COVID-19 risk yet increase the susceptibility for allergic asthma, this study exposes a bidirectional axis of evolutionary trade-offs that mirrors the HDCP paradigm: the same immune phenotype that protects against acute infection could, when misdirected, precipitate chronic inflammatory disease.

Together, these observations imply that infection-responsive immune circuits should be mapped as dynamic traits that traverse a spectrum from protective host defense to pathology. Incorporating allergic phenotypes into HDCP-style multi-omic atlases will therefore be critical for identifying the immunogenetic “tipping points” at which beneficial barrier immunity yields to maladaptive disease.

## Declarations

### Ethical Approval and Consent to participate

No human subjects were directly involved in this study. All the data used in this study was derived from existing de-identified biological samples from prior studies. Thus, ethical and patient consent was not required in this study.

### Competing interests

The funders had no role in the study design, data analysis, data interpretation, and writing of this manuscript. This study was conducted in the absence of any commercial or financial relationships that could be construed as a potential conflict of interest.

## Data Availability

All data produced in the present study are available upon reasonable request to the authors.

## Acknowledgements

We would like to acknowledge the participants and investigators of the COVID-19 Host Genetics Initiative, the FinnGen project, and the UK biobank.

## Availability of supporting data

The data that support the findings of this study will be available from the corresponding author upon reasonable request.

## Funding

This work is primarily supported by the Human Disease Continuum Project (HDCP) from M.Z.’s personal funds and resources. All funding sources have been disclosed, and no additional financial support was received for this study.

## Authors’ contributions

M.Z. conceived the project, designed the study, developed the method, conducted data analysis, and wrote the manuscript. M.Z. supervised this project and is responsible for the overall content.

## Nonstandard Abbreviations and Acronyms

COVID-19: coronavirus disease 2019
MR: Mendelian randomization
MR-PRESSO: Mendelian randomization pleiotropy residual sum and outlier
GWAS: genome-wide association study
SNP: single nucleotide polymorphism
LD: linkage disequilibrium

---

**Supplementary Figure 1.**
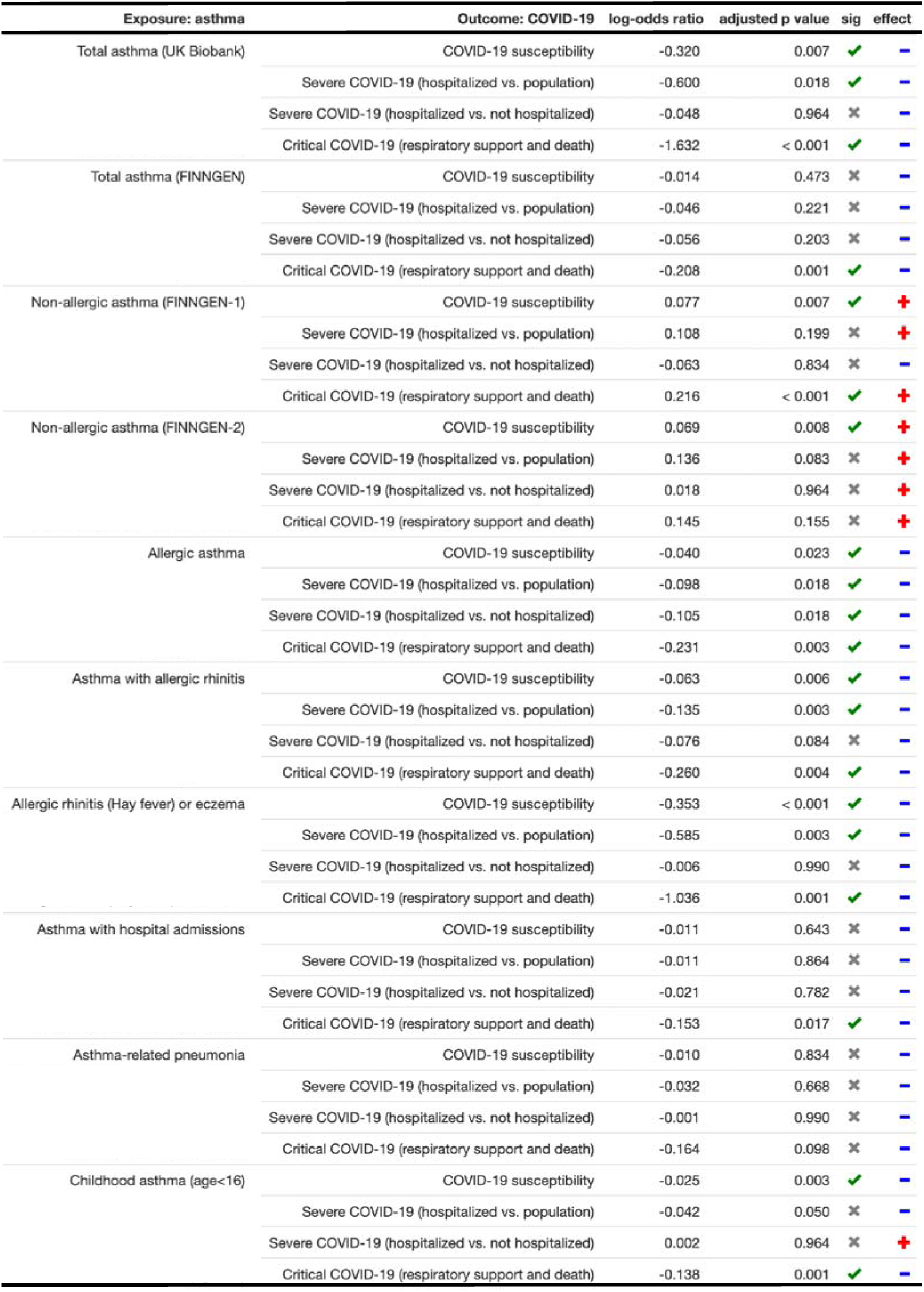
Mendelian randomization analysis of asthma and COVID-19.

## Notes

### Competing Interest Statement

The authors have declared no competing interest.

### Funding Statement

This work is primarily supported by the Human Disease Continuum Project (HDCP) from M.Z. personal funds and resources. All funding sources have been disclosed, and no additional financial support was received for this study.

