## Supplemental Figure. 1 for "Allergic asthma and type-2 immunity reduce COVID-19 severity"

| Exposure: asthma | Outcome: COVID-19 | log-odds ratio | adjusted p value | sig | effect |
|---|---|---|---|---|---|
| Total asthma (UK Biobank) | COVID-19 susceptibility | -0.320 | 0.007 | ✓ | − |
| | Severe COVID-19 (hospitalized vs. population) | -0.600 | 0.018 | ✓ | − |
| | Severe COVID-19 (hospitalized vs. not hospitalized) | -0.048 | 0.964 | ✗ | − |
| | Critical COVID-19 (respiratory support and death) | -1.632 | < 0.001 | ✓ | − |
| Total asthma (FINNGEN) | COVID-19 susceptibility | -0.014 | 0.473 | ✗ | − |
| | Severe COVID-19 (hospitalized vs. population) | -0.046 | 0.221 | ✗ | − |
| | Severe COVID-19 (hospitalized vs. not hospitalized) | -0.056 | 0.203 | ✗ | − |
| | Critical COVID-19 (respiratory support and death) | -0.208 | 0.001 | ✓ | − |
| Non-allergic asthma (FINNGEN-1) | COVID-19 susceptibility | 0.077 | 0.007 | ✓ | + |
| | Severe COVID-19 (hospitalized vs. population) | 0.108 | 0.199 | ✗ | + |
| | Severe COVID-19 (hospitalized vs. not hospitalized) | -0.063 | 0.834 | ✗ | − |
| | Critical COVID-19 (respiratory support and death) | 0.216 | < 0.001 | ✓ | + |
| Non-allergic asthma (FINNGEN-2) | COVID-19 susceptibility | 0.069 | 0.008 | ✓ | + |
| | Severe COVID-19 (hospitalized vs. population) | 0.136 | 0.083 | ✗ | + |
| | Severe COVID-19 (hospitalized vs. not hospitalized) | 0.018 | 0.964 | ✗ | + |
| | Critical COVID-19 (respiratory support and death) | 0.145 | 0.155 | ✗ | + |
| Allergic asthma | COVID-19 susceptibility | -0.040 | 0.023 | ✓ | − |
| | Severe COVID-19 (hospitalized vs. population) | -0.098 | 0.018 | ✓ | − |
| | Severe COVID-19 (hospitalized vs. not hospitalized) | -0.105 | 0.018 | ✓ | − |
| | Critical COVID-19 (respiratory support and death) | -0.231 | 0.003 | ✓ | − |
| Asthma with allergic rhinitis | COVID-19 susceptibility | -0.063 | 0.006 | ✓ | − |
| | Severe COVID-19 (hospitalized vs. population) | -0.135 | 0.003 | ✓ | − |
| | Severe COVID-19 (hospitalized vs. not hospitalized) | -0.076 | 0.084 | ✗ | − |
| | Critical COVID-19 (respiratory support and death) | -0.260 | 0.004 | ✓ | − |
| Allergic rhinitis (Hay fever) or eczema | COVID-19 susceptibility | -0.353 | < 0.001 | ✓ | − |
| | Severe COVID-19 (hospitalized vs. population) | -0.585 | 0.003 | ✓ | − |
| | Severe COVID-19 (hospitalized vs. not hospitalized) | -0.006 | 0.990 | ✗ | − |
| | Critical COVID-19 (respiratory support and death) | -1.036 | 0.001 | ✓ | − |
| Asthma with hospital admissions | COVID-19 susceptibility | -0.011 | 0.643 | ✗ | − |
| | Severe COVID-19 (hospitalized vs. population) | -0.011 | 0.864 | ✗ | − |
| | Severe COVID-19 (hospitalized vs. not hospitalized) | -0.021 | 0.782 | ✗ | − |
| | Critical COVID-19 (respiratory support and death) | -0.153 | 0.017 | ✓ | − |
| Asthma-related pneumonia | COVID-19 susceptibility | -0.010 | 0.834 | ✗ | − |
| | Severe COVID-19 (hospitalized vs. population) | -0.032 | 0.668 | ✗ | − |
| | Severe COVID-19 (hospitalized vs. not hospitalized) | -0.001 | 0.990 | ✗ | − |
| | Critical COVID-19 (respiratory support and death) | -0.164 | 0.098 | ✗ | − |
| Childhood asthma (age<16) | COVID-19 susceptibility | -0.025 | 0.003 | ✓ | − |
| | Severe COVID-19 (hospitalized vs. population) | -0.042 | 0.050 | ✗ | − |
| | Severe COVID-19 (hospitalized vs. not hospitalized) | 0.002 | 0.964 | ✗ | + |
| | Critical COVID-19 (respiratory support and death) | -0.138 | 0.001 | ✓ | − |
